# Atrial Fibrillation Inpatient Management Patterns and Clinical Outcomes During The Conflict In Syria: An Observational Cohort Study

**DOI:** 10.1101/2023.09.04.23294670

**Authors:** Ibrahim Antoun, Alkassem Alkhayer, Ahmed Kotb, Ibrahim Hanafi, Yaman Mahfoud, Joseph Barker, Akash Mavilakandy, Alamer Alkhayer, Saif Shah, Peter Simon, Riyaz Somani, G Andre’ Ng

**Author notes:** Corresponding author Ibrahim Antoun MD MRCP PhD, Department of Cardiovascular Sciences, Glenfield Hospital | Groby Road | Leicester | LE3 9QP | UK.

## Abstract

**Background:** Atrial fibrillation (AF) is the most common sustained arrhythmia worldwide. However, there is no data on AF inpatient management strategies and clinical outcomes in Syria.

**Objectives:** This study aims to navigate AF inpatient management and cardiovascular (CV) mortality in a tertiary cardiology centre in Latakia, Syria.

**Methods:** A single-centre retrospective observational cohort study was conducted at Tishreen’s University Hospital, Latakia, Syria, from June 2021 to June 2023. We included patients ≥16 years of age presenting and being treated for AF as the primary diagnosis at the emergency department. Medical records were examined for patient demographics, laboratory results, treatment plans and inpatient details. Inpatient cardiovascular mortality was the primary outcome of interest.

**Results:** The study included 596 patients. The median age was 58 and 61% were males. Only 121 patients (20.3%) were known to have AF. Rhythm control strategy was persuaded in 39% of patients, with Amiodarone being the only agent utilised while the rest adopted a rate control approach. CV Mortality occurred in 28 patients (4.7%). The presence of smoking (aOR: 1.6, 95% CI: 1.3 to 2.1, P=0.04), thyroid disease (aOR= 9.7, 95% CI= 1.2 to 91.6, P<0.001), COPD (aOR: 82, 95% CI: 12.7 to 711, P<0.001) and valvular heart disease (VHD) (aOR= 9.1, 95% CI: 1.7 to 55.1, P<0.001) were independent risk factors of cardiovascular inpatient mortality.

**Conclusion:** Current smoking, thyroid disease, COPD and VHD were independent risk factors of inpatient cardiovascular mortality with AF in Syria. Elective rhythm control was challenging due to limited resources.

## Background

Atrial fibrillation (AF) is the most common sustained arrhythmia worldwide, and prevalence in low to middle-income countries is likely underestimated (1). Although AF in the Western world is extensively studied, little data corroborates AF demographics and management in the Middle East, with only four epidemiological data registries (2). AF-related research is still insufficient in the Arab world, contributing only 0.7% of the total AF research (3).

One of these countries, Syria, has been in conflict since 2011 and deprived of healthcare resources and funding, particularly exacerbated during the COVID-19 pandemic. As a result, less than half of its hospitals operate at full performance, with over 50% of its healthcare workforce forced to leave due to conflict. More than half the Syrian population live in poverty, which is suggested to increase AF risk (4). The AF management in hospitals during the accompanying political and economic turmoil is unclear, with a lack of published inpatient outcomes and figures originating from Syrian hospitals. In the context of the constraint resources, a real-world depiction of currently practised AF care and observed outcomes could potentially aid management and allocation of resources by identifying remediable areas of deficiencies and, more importantly, reasonable and practical solutions that could be implemented.

We aimed to describe the characteristics and outcomes of patients treated for AF in Latakia’s cardiology tertiary care centre over two years.

## Methods

This is a single-centre retrospective observational cohort study conducted at Tishreen’s University Hospital, Latakia, Syria, between the 1^st^ of June 2021 and the 1^st^ of June 2023. Ethical approval was obtained by the hospital board and the ethical board at Tishreen’s University Hospital (reference: 277/A).

Inclusion criteria were adult patients (over the age of 16 years) presenting to the hospital and treated for AF as the primary diagnosis. Patients exhibiting AF as a secondary component or complication to another primary diagnosis were excluded. The diagnosis of AF was established by 12-lead electrocardiograms (ECG) and subsequently interpreted and verified by a consultant cardiologist.

Patient demographics, admission details, 12-lead ECGs, blood results, radiological reports and clinical outcomes during hospitalisation were extracted from patients’ records. The primary outcomes included the length of hospital stay and in-hospital cardiovascular (CV) mortality. The study was written according to STROBE guidelines — see Supplementary Table 1 (5).

### Statistical analysis

Continuous variables are expressed as median and interquartile ranges (IQR). Categorical variables are expressed as counts and percentages (%). Pearson’s χ 2 or Fisher’s exact test was used for categorical variables between groups. Student’s *t-*test and Kruskal-Wallis test were used to compare continuous variables between the groups depending on the normality of the distribution.

Multiple logistic regression models were used to investigate the relationship between variables and mortality. We hypothesised that specific demographic characteristics and comorbidities would affect AF mortality. Therefore, we constructed a base model consisting of age and gender to assess the incremental value of comorbidities that are significantly associated with mortality. These comorbidities were then added to the base model to improve the mortality predictability of the model. The new model’s cumulative discrimination was measured using the area under the curve; its statistical significance compared to the base model was assessed with the likelihood ratio test. A 2-sided p-value <0.05 was considered statistically significant. Statistical analysis was performed using GraphPad Prism V10.0 for Mac (San Diego, California, USA; www.graphpad.com).

## Results

### Baseline characteristics, presentation, and diagnostics tests

Between the 1^st^ of June 2021 and the 1^st^ of June 2023, 596 patients presented to the hospital with AF as the primary diagnosis on admission, all of whom were of Arab descent. Only 121 patients (20%) were known to have AF before presentation without the ability to differentiate between paroxysmal and persistent AF due to the lack of Holter monitoring and previous clinic follow-up. The most common presenting complaint was palpitations in 335 patients (56%), followed by shortness of breath in 171 patients (29%), chest pain in 40 patients (7%), clinical heart failure in 28 patients (5%) and syncope in 23 patients (4%). Around three-quarters of patients were current smokers. Demographic and lab results are demonstrated in Table 1. Moderate to severe valvular heart disease (VHD) was present in 90 patients (15%), of which 20 (22%) had rheumatic mitral stenosis, 18 (20%) had moderate to severe tricuspid regurgitation, 29 (21%) had moderate to severe mitral regurgitation, 30 (33%) had moderate to severe aortic stenosis, and three (3%) had surgically corrected metallic aortic valve. This information was extracted from previous echocardiograms that patients underwent. Only 3 out of 20 patients (15%) with active thyroid disease were on medical treatment. When stratified by length Of stay, patients admitted for ≥ four days had higher rates of smoking, hypertension, VHD, and faster heart rate (HR) on admission.

### Treatments

Management strategies were stratified into rhythm control, rate control or combined approach based on clinical judgement and available resources. A rhythm control strategy was adopted in 230 patients (39%). This was done exclusively using pharmacological therapy, with amiodarone being the only drug available. The regime used for all patients was an intravenous (IV) loading dose of 300mg followed by an oral maintenance dose of 200 mg twice a day (BD) for a week, then 200 mg once a day (OD) after that. Bisoprolol was used with amiodarone for additional rate control in 131 patients (57% of amiodarone patients). Direct current cardioversion (DCCV) was used in an emergency setting for ten patients during admission due to hemodynamic instability, with eight being successful after a single shock and two passing away due to degeneration to asystole. Elective DCCV was not used in this cohort, nor were any patients scheduled for one after discharge. None of the patients was referred to catheter ablation or left atrial appendage closure, which is not offered in the city. Monotherapy rate control was employed in 366 patients (61%). Metoprolol was the only IV rate-control drug used in 90 patients (15%) with 5 mg to 15 mg boluses. The two available oral drugs were bisoprolol 2.5-10 mg, used in 249 patients (42%), followed by verapamil 120mg BD to three times a day (TDS), used in 28 patients (5%). The HR was reduced significantly in all patients on discharge from 137 (120-152) BPM to 76 (65-90) BPM, P<0.0001.

Oral anticoagulation was used in 500 patients (84%) based on the CHA_2_DS_2_-VASc score (cut-offs of ≥1 for males and ≥2 for females were used for anticoagulation) or if a rhythm control strategy was adopted. Of these, Warfarin was used in 21 patients (4%), of which 18 had rheumatic heart disease, and three had a metallic heart valve. The rest of the patients (n=479) received a direct oral anticoagulant (DOAC) based on availability. The available DOACs were apixaban (2.5-5mg BD) and rivaroxaban (15-20mg OD). Urological and gastrointestinal bleeds occurred in 12 patients within 36 hours of anticoagulation initiation, but none were fatal. Eight patients resumed lower-dose DOAC after careful patient counselling and shared decision-making, while the rest discontinued, accepting the risk of cardio-embolic events.

### Inpatient outcomes and predictors

Inpatient mortality was 5% (n=31) after a median hospital admission of 4 (3-7) days. CV mortality occurred in 28 patients (4.7%), while three died from hospital-acquired infections (0.3%). The remaining patients were discharged home after a median of 4 (2-5) days. Limited facilities and recourses did not allow patients to be scheduled for an outpatient follow-up clinic or diagnostic tests such as a Holter monitor or a transthoracic echocardiogram. Table 2 demonstrates univariable and multivariable logistic regressions in relation to mortality. The univariable logistic regression model concluded that valvular heart disease, COPD, smoking, IHD and thyroid disease with a higher T4 were associated with increased inpatient mortality. In a multivariable model consisting of age, gender, smoking status, thyroid disease, COPD and VHD, it was found that the presence of smoking (aOR: 1.6, 95% CI: 1.3 to 2.1, P=0.04, thyroid disease (aOR= 9.7, 95% CI= 1.2 to 91.6, P<0.001), COPD (aOR: 82, 95% CI: 12.7 to 711, P<0.001) and VHD (aOR= 9.1, 95% CI: 1.7 to 55.1, P<0.001) were independent risk factors of CV inpatient mortality. Adding smoking, COPD, thyroid disease, and VHD to the base model provided the best AUC to predict inpatient mortality (AUC=0.9, Table 3.

## Discussion

This is the first study to address AF inpatient treatment patterns and clinical outcomes in Syria, a country suffering from conflict since 2011 and recently hit by devastating earthquakes (6). We report an AF inpatient cohort of 596 patients. The main findings include that the hospitalised AF Syrian patients were relatively younger than other neighbouring countries (7), the lack of elective rhythm control regardless of patient’s age, comorbidities or symptom burden due to the paucity of resources, so there was blank rate control strategy adopted in most patients, and that current smoking, thyroid disease, COPD and VHD were independent risk factors of CV inpatient mortality in a prediction model that has an AUC of 0.9.

There has been a significant increase in AF prevalence in low-income countries in the last 20 years (8). The Syrian population have been living in poverty and life stress for the previous ten years, both of which are suggested to promote AF (4, 9). This inpatient cohort is relatively younger than patients admitted in high-income countries. For example, 453,060 hospitalised patients with AF in the US had a mean age of 70.1, more than our cohort (10). Our cohort was even younger than the Jordanian cohort, with a mean age of 68 (7). This is likely due to the lack of primary or secondary care risk factor control owing to the lack of funding and resources. The high smoking rate likely affected our cohort’s AF incidence (11).

Moreover, clinicians did adopt the latest evidence and treatment guidelines to the best of their abilities, but limited resources hindered this (12). Appropriate rate and rhythm control strategies were conducted using available resources, and DCCV was appropriately used in hemodynamically unstable patients. This contributed to the relatively acceptable inpatient outcomes, knowing the challenging circumstances in Syria. There is ongoing evidence worldwide about the benefit of early rhythm control on symptoms, stroke risk, quality of life and cardiovascular outcomes, which can benefit this cohort (13-15). However, this approach could not be implemented due to the intensive follow-up, resources and the availability of antiarrhythmic drugs and ablation centres, which are unavailable in Syria. (16). This should be an area for improvement to optimise long-term outcomes and avoid re-admissions. Therefore, follow-up data is suggested in future studies.

Our model was highly predictive of inpatient CV mortality. Current smoking (17) and COPD (18) were associated with increased mortality in previous studies but not in a predominantly Arab cohort. This study could be used to validate these findings in such ethnicity. A history of thyroid disease is known to increase AF risk but has not been shown to increase mortality (19). Almost 15% of our cohort with thyroid disease were on active treatment, which could have contributed to current results. To optimise AF treatment, active steps should be taken to educate patients about modifiable risk factors and lifestyle triggers. VHD is known to cause AF (20), and a community-based study suggested that moderate to severe biological VHD increased mortality (21). This can be reflected in our cohort, especially with the difficulty of accessing a cardiac surgery service due to poor/damaged infrastructure. Many of these patients remain untreated, which contributes to increased mortality. This highly predictive model can be used from a patient’s perspective to address smoking (which will improve COPD management) and encourage compliance with thyroid medications.

Furthermore, the predictive model could be essential for policymakers to ensure these patients have follow-up to address risk factors, especially a potential need for surgical treatment for VHD. From a physician’s perspective, this model helps identify patients at a high risk of deterioration to ensure early escalation and prompt treatment. In case of limited follow-up availability, these patients could be prioritised to prevent CV mortality. More resources are required to ensure guidelines adherence and follow-up of the most vulnerable patients to achieve better outcomes.

## Conclusion

Syrian inpatients admitted acutely with AF are relatively young, and the limited resources prevented optimal AF management, especially elective rhythm control, despite physicians’ best efforts. Current smoking, thyroid disease, COPD and VHD were independent risk factors for CV inpatient mortality.

### Limitations

Our study had limitations. Our data collection was limited to a single tertiary care centre in Latakia. This city was less affected by the Syrian war than other northern and eastern regions of Syria. Therefore, our results might not generalise to other centres/regions, knowing the extreme heterogeneity in the quality and quantity of hospital supplies and staff. Additionally, our analysis included only routinely collected data within the medical records and by the number of patients who presented to the hospital. Therefore, we missed the other variables that may have impacted mortality. Nevertheless, our study successfully provided significant, timely findings that may prove helpful in improving the care provided and reducing mortality in patients with AF.

## Data Availability

Available upon request of the corresponding author

## Data availability

Data relating to this study are available upon reasonable request from the corresponding author.

## Acknowledgements

AK is supported by a clinical research fellowship from Abbott. GA Ng is supported by a British Heart Foundation Programme Grant (RG/17/3/32,774) and the Medical Research Council Biomedical Catalyst Developmental Pathway Funding Scheme (MR/S037306/1). JB is supported by the NIHR Academic Clinical Fellowship.

## Ethical statement

This study was reviewed and approved by the institutional ethical committee of the Tishreen University Hospital.

## Authors contributions

IA designed the study, analysed the data and wrote the first manuscript draft. AA, AA, AT and YM managed data collection. AK, IH, SS, AM, GAN, RS, and JB reviewed and edited the manuscript.

## References

1. Rahman F, Kwan GF, Benjamin EJ. Global epidemiology of atrial fibrillation. Nature Reviews Cardiology. 2014;11(11):639.

2. Al-Shamkhani W, Ayetey H, Lip GY. Atrial fibrillation in the Middle East: unmapped, underdiagnosed, undertreated. AtrialExpert review of cardiovascular therapy. 2018;16(5):341–8.

3. Akiki D, El Hage S, Wakim E, Safi S, Assouad E, Salameh P. Atrial Fibrillation in the Arab World: A Bibliometric Analysis of Research Activity from 2004 to 2019. AtrialJournal of Cardiac Arrhythmias. 2021;34(1):12–22.

4. Essien UR, McCabe ME, Kershaw KN, Youmans QR, Fine MJ, Yancy CW, et al. Association Between Neighborhood-Level Poverty and Incident Atrial Fibrillation: a Retrospective Cohort Study. AtrialJ Gen Intern Med. 2022;37(6):1436–43.

5. Vandenbroucke JP, Von Elm E, Altman DG, Gøtzsche PC, Mulrow CD, Pocock SJ, et al. Strengthening the Reporting of Observational Studies in Epidemiology (STROBE): explanation and elaboration. AtrialPLoS medicine. 2007;4(10):e297.

6. Naddaf M. Turkey-Syria earthquake: What scientists know. AtrialNature. 2023.

7. Hammoudeh A, Khader Y, Tabbalat R, Badaineh Y, Kadri N, Shawer H, et al. One-Year Clinical Outcome in Middle Eastern Patients with Atrial Fibrillation: The Jordan Atrial Fibrillation (JoFib) Study. AtrialInternational Journal of Vascular Medicine. 2022;2022:4240999.

8. Lippi G, Sanchis-Gomar F, Cervellin G. Global epidemiology of atrial fibrillation: an increasing epidemic and public health challenge. AtrialInternational Journal of Stroke. 2020:1747493019897870.

9. Patel D, Mc Conkey ND, Sohaney R, Mc Neil A, Jedrzejczyk A, Armaganijan L. A systematic review of depression and anxiety in patients with atrial fibrillation: the mind-heart link. AtrialCardiovascular Psychiatry and Neurology. 2013;2013.

10. Jackson SL, Tong X, Yin X, George MG, Ritchey MD. Emergency Department, Hospital Inpatient, and Mortality Burden of Atrial Fibrillation in the United States, 2006 to 2014. AtrialThe American Journal of Cardiology. 2017;120(11):1966–73.

11. Aune D, Schlesinger S, Norat T, Riboli E. Tobacco smoking and the risk of atrial fibrillation: a systematic review and meta-analysis of prospective studies. AtrialEuropean journal of preventive cardiology. 2018;25(13):1437–51.

12. Hindricks G, Potpara T, Dagres N, Arbelo E, Bax JJ, Blomström-Lundqvist C, et al. 2020 ESC Guidelines for the diagnosis and management of atrial fibrillation developed in collaboration with the European Association for Cardio-Thoracic Surgery (EACTS): The Task Force for the diagnosis and management of atrial fibrillation of the European Society of Cardiology (ESC) Developed with the special contribution of the European Heart Rhythm Association (EHRA) of the ESC. AtrialEuropean Heart Journal. 2020;42(5):373–498.

13. Kirchhof P, Camm AJ, Goette A, Brandes A, Eckardt L, Elvan A, et al. Early Rhythm-Control Therapy in Patients with Atrial Fibrillation. AtrialNew England Journal of Medicine. 2020;383(14):1305–16.

14. Guertin JR, Dorais M, Khairy P, Sauriol L, Matteau A, Poulin F, et al. Atrial fibrillation: a reallife observational study in the Québec population. AtrialCan J Cardiol. 2011;27(6):794–9.

15. Kim D, Yang P-S, You SC, Sung J-H, Jang E, Yu HT, et al. Treatment timing and the effects of rhythm control strategy in patients with atrial fibrillation: nationwide cohort study. Atrialbmj. 2021;373.

16. Fouad FM, Sparrow A, Tarakji A, Alameddine M, El-Jardali F, Coutts AP, et al. Health workers and the weaponisation of health care in Syria: a preliminary inquiry for The Lancet–American University of Beirut Commission on Syria. AtrialThe Lancet. 2017;390(10111):2516–26.

17. Albertsen IE, Rasmussen LH, Lane DA, Overvad TF, Skjøth F, Overvad K, et al. The Impact of Smoking on Thromboembolism and Mortality in Patients With Incident Atrial Fibrillation. AtrialChest. 2014;145(3):559–66.

18. Ye J, Yao P, Shi X, Yu X. A systematic literature review and meta-analysis on the impact of COPD on atrial fibrillation patient outcome. AtrialHeart & Lung. 2022;51:67–74.

19. Bruere H, Fauchier L, Brunet AB, Pierre B, Simeon E, Babuty D, et al. History of thyroid disorders in relation to clinical outcomes in atrial fibrillation. AtrialThe American Journal of Medicine. 2015;128(1):30–7.

20. Benjamin EJ, Levy D, Vaziri SM, D’Agostino RB, Belanger AJ, Wolf PA. Independent risk factors for atrial fibrillation in a population-based cohort. AtrialThe Framingham Heart Study. Jama. 1994;271(11):840–4.

21. Thomas KL, Jackson LR, Shrader P, Ansell J, Fonarow GC, Gersh B, et al. Prevalence, characteristics, and outcomes of valvular heart disease in patients with atrial fibrillation: insights from the ORBIT-AF (outcomes registry for better informed treatment for atrial fibrillation). AtrialJournal of the American Heart Association. 2017;6(12):e006475.

