## Supplementary material for "Atrial Fibrillation Inpatient Management Patterns and Clinical Outcomes During The Conflict In Syria: An Observational Cohort Study": Tables

Table 1: Patient details, admission, vital signs and lab workup stratified by admission time

| Variable | Full cohort (n=596) | ≥4 days (n=315) | 4< days (n=281) | p-value |
| --- | --- | --- | --- | --- |
| Age (years) | 58 (51-65) | 55 (48-63) | 60 (54-67) | 0.19 |
| Male | 366 (61%) | 211 (67%) | 155 (56%) | 0.14 |
| Smoking | 232 (74%) | 188 (74%) | 66 (24%) | <0.0001 |
| Hypertension | 220 (37%) | 132 (42%) | 88 (32%) | 0.001 |
| Ischemic heart disease | 101 (17%) | 53 (17%) | 58 (21%) | 0.4 |
| Diabetes mellitus | 136 (23%) | 69 (22%) | 67 (24%) | 0.39 |
| Cerebrovascular disease | 108 (18%) | 51 (16%) | 57 (20%) | 0.1 |
| Chronic obstructive lung disease | 51 (9%) | 21 (8%) | 30 (11%) | 0.2 |
| Heart failure | 108 (18%) | 53 (17%) | 65 (23%) | 0.1 |
| Thyroid disease | 20 (3%) | 17 (5%) | 3 (1%) | 0.015 |
| CHA_2_DS_2_-VASc | 2 (1-3) | 2 (1-3) | 2 (1-3) | 0.15 |
| Valvular heart disease | 90 (15%) | 56 (18%) | 34 (12%) | <0.0001 |
| Haemoglobin (g/dL) | 11 (9-13) | 10 (9-13) | 11 (10-14) | 0.29 |
| C-reactive protein (mg/dL) | 40 (22-65) | 39 (22-65) | 43 (23-67) | 0.71 |
| White cell count (10^9^/L) | 9 (6-12) | 8 (5-13) | 9 (5-12) | 0.92 |
| Rate on admission (beats per minute) | 137 (120-152) | 139 (124-155) | 135 (117-150) | 0.005 |
| Rate on discharge (beats per minute) | 76 (65-90) | 75 (66-90) | 77 (66-89) | 0.73 |
| T4 (ug/dL) | 9.6 (7.1-13) | 9.4 (6.9-12) | 9.4 (6.7-13) | 0.85 |
| Thyroid stimulating hormone (mlUl/L) | 2.3 (1.3-3.2) | 2.2 (1.2-33.2) | 2.5 (1.4-3.3) | 0.26 |
| Magnesium (mmol/L) | 0.9 (0.6-1.1) | 0.8 (0.6-1.1) | 0.9 (0.6-1.1) | 0.8 |
| Potassium (mmol/L) | 4 (.4-4.6) | 4.1 (3.4-4.7) | 4 (3.5-4.6) | 0.76 |
| Calcium (mmol/L) | 2.2 (2.1-2.4) | 2.3 (2.2-2.5) | 2.2 (2.1-2.3) | 0.85 |

Table 2: Univariable and multivariable logistic regressions in relation to inpatient mortality. Variables that showed a statistically significant correlation with length of stay in the univariable analysis were included in the multivariable analysis, along with age and sex (base model).

|  | Univariable analysis | | |  | | Multivariable analysis | |
| --- | --- | --- | --- | --- | --- | --- | --- |
| Variables | OR | CI | p-value | | aOR | CI | p-value |
| Age (per year increase) | 1.1 | 1.0 to 1.1 | 0.06 | | 1.1 | 0.9 to 1.2 | 0.09 |
| Sex (male compared to female) | 1.4 | 0.62 to 3.1 | 0.41 | | 2.2 | 0.3 to 16.6 | 0.4 |
| Hypertension (yes vs no) | 0.9 | 0.38 to 2.0 | 0.8 | |  |  |  |
| Diabetes mellitus (yes vs no) | 0.76 | 0.10 to 1.3 | 0.17 | |  |  |  |
| Cerebrovascular event (yes vs no) | 0.77 | 0.13 to 1.7 | 0.37 | |  |  |  |
| Ischemic heart disease | 3.3 | 0.4 to 7.6 | 0.047 | |  |  |  |
| Heart failure (yes vs no) | 1.4 | 0.54 to 3.3 | 0.44 | |  |  |  |
| Valvular heart disease (yes vs no) | 13 | 5.8 to 33 | <0.001 | | 9.1 | 1.7 to 55.1 | <0.001 |
| Chronic obstructive lung disease (yes vs no) | 98 | 34 to 355 | <0.001 | | 82.0 | 12.7 to 711 | <0.001 |
| CHA2S2Vasc score (per 1 point increase) | 1.1 | 0.88 to 1.5 | 0.31 | |  |  |  |
| Thyroid disease (yes vs no) | 49 | 15 to 176 | <0.001 | | 9.7 | 1.2 to 91.6 | <0.001 |
| Smoking (yes vs no) | 2.5 | 1.0 to 6.9 | 0.054 | | 1.6 | 1.3 to 2.1 | 0.04 |
| White cell count (per unit-10^9^/L increase) | 1.1 | 0.95 to 1.2 | 0.32 | |  |  |  |
| Haemoglobin (per unit- g/dL increase) | 1.2 | 1.0 to 1.4 | 0.09 | |  |  |  |
| C-reactive protein (per unit- mg/dL increase) | 1 | 0.99 to 1.0 | 0.34 | |  |  |  |
| Calcium (per unit- mmol/L increase) | 0.65 | 0.059 to 13 | 0.07 | |  |  |  |
| T4 (per unit- mmol/L increase) | 1.4 | 1.2 to 1.6 | 0.001 | |  |  |  |
| TSH (per unit- mmol/L increase) | 0.45 | 0.32 to 0.72 | 0.05 | |  |  |  |
| Magnesium (per unit- mmol/L increase) | 1.5 | 0.39 to 6.0 | 0.5 | |  |  |  |
| Potassium (per unit- mmol/L increase) | 0.95 | 0.57 to 1.6 | 0.84 | |  |  |  |
| Heart rate (per beat per minute increase) | 1 | 0.99 to 1.0 | 0.35 | |  |  |  |

Table 3 Comparison between different multivariable logistic regressions in relation to

inpatient mortality

| Model | AUC | P |
| --- | --- | --- |
| Base model | 0.59 | 0.12 |
| +IHD | 0.61 | 0.051 |
| +VHD | 0.81 | 0.001 |
| +Thyroid disease, T4, TSH, | 0.85 | 0.002 |
| +COPD | 0.86 | 0.001 |
| +VHD and COPD | 0.87 | 0.001 |
| +Smoking, COPD, VHD | 0.87 | 0.001 |
| +Smoking, thyroid disease, COPD, VHD | 0.9 | <0.001 |
